# SARS-CoV-2 evolved during advanced HIV disease immunosuppression has Beta-like escape of vaccine and Delta infection elicited immunity

**DOI:** 10.1101/2021.09.14.21263564

**Authors:** Sandile Cele, Farina Karim, Gila Lustig, James Emmanuel San, Tandile Hermanus, Houriiyah Tegally, Jumari Snyman, Thandeka Moyo-Gwete, Eduan Wilkinson, Mallory Bernstein, Khadija Khan, Shi-Hsia Hwa, Sasha W. Tilles, Lavanya Singh, Jennifer Giandhari, Ntombifuthi Mthabela, Matilda Mazibuko, Yashica Ganga, Bernadett I. Gosnell, Salim Abdool Karim, Willem Hanekom, Wesley C. Van Voorhis, Thumbi Ndung’u, COMMIT-KZN Team, Richard J. Lessells, Penny L. Moore, Mahomed-Yunus S. Moosa, Tulio de Oliveira, Alex Sigal

## Abstract

Characterizing SARS-CoV-2 evolution in specific geographies may help predict the properties of variants coming from these regions. We mapped neutralization of a SARS-CoV-2 strain that evolved over 6 months from the ancestral virus in a person with advanced HIV disease. Infection was before the emergence of the Beta variant first identified in South Africa, and the Delta variant. We compared early and late evolved virus to the ancestral, Beta, Alpha, and Delta viruses and tested against convalescent plasma from ancestral, Beta, and Delta infections. Early virus was similar to ancestral, whereas late virus was similar to Beta, exhibiting vaccine escape and, despite pre-dating Delta, strong escape of Delta-elicited neutralization. This example is consistent with the notion that variants arising in immune-compromised hosts, including those with advanced HIV disease, may evolve immune escape of vaccines and enhanced escape of Delta immunity, with implications for vaccine breakthrough and reinfections.

**Highlights:**

- A prolonged ancestral SARS-CoV-2 infection pre-dating the emergence of Beta and Delta resulted in evolution of a Beta-like serological phenotype
- Serological phenotype includes strong escape from Delta infection elicited immunity, intermediate escape from ancestral virus immunity, and weak escape from Beta immunity
- Evolved virus showed substantial but incomplete escape from antibodies elicited by BNT162b2 vaccination

**Graphical abstract:** 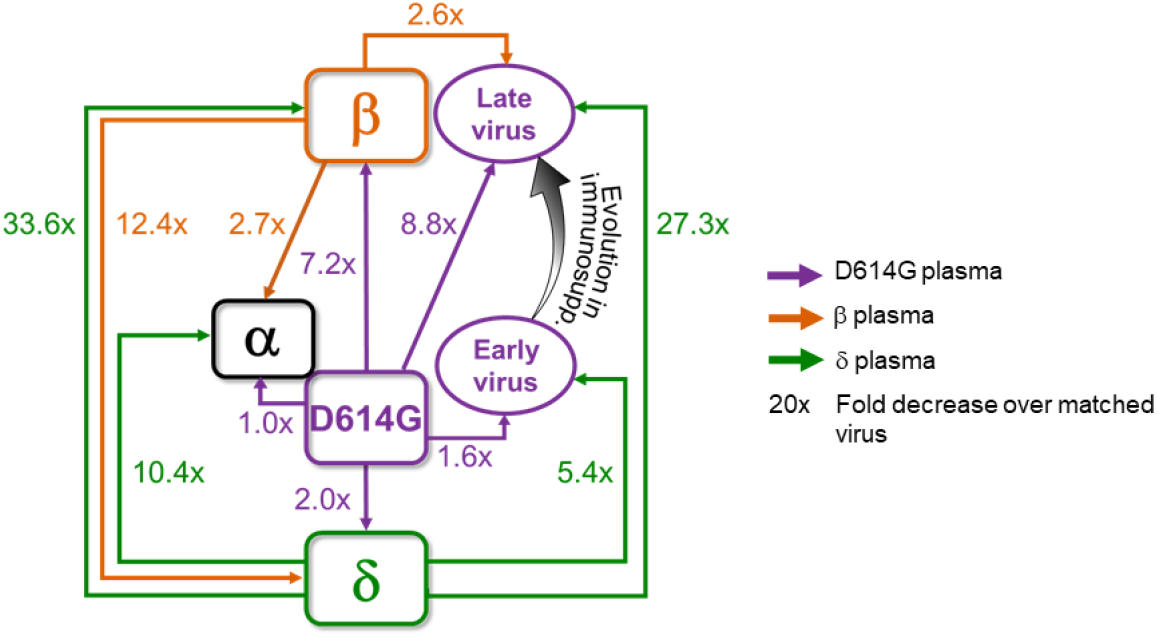

---

Neutralization is highly predictive of vaccine efficacy (1, 2). Some SARS-CoV-2 variants show decreased neutralization by vaccine elicited immunity. This may make vaccines less effective at reducing the frequency of infection (3). The Alpha variant shows relatively little escape (4-7). The Beta (4, 6-12), Gamma (13), Lambda (14) and Mu (15) variants show neutralization escape to different degrees from neutralization elicited by ancestral SARS-CoV-2 infection or vaccines. The Delta variant has evolved a strong transmission advantage over other SARS-CoV-2 strains (16). It does not show a high degree of neutralization escape from plasma immunity elicited by ancestral strains but does show escape from Beta antibody immunity (17).

Escape from neutralization and enhanced transmission involves substitutions and deletions in the spike glycoprotein of the virus which binds the ACE2 receptor on the cell surface (18). Mutations associated with neutralization escape are found in the receptor binding domain (RBD) (18-20), and N-terminal domain of spike (21-24). RBD mutations for the Beta variant include the K417N, E484K and N501Y (see https://covdb.stanford.edu/page/mutation-viewer/). E484K and N501Y are shared with Gamma, which has the K417T instead of K417N. Alpha shares N501Y and in some cases E484K. Lambda has L452Q and F490S. Delta has L452R and T478K. There are also extensive differences in the NTD. For example, Beta NTD substitutions include L18F, D80A, D215G, and a 241-243 deletion. In contrast, Delta has T19R, G142D, E156G, and a 157-158 deletion.

An important consideration in some geographical areas is high prevalence of co-infection of SARS-CoV-2 and HIV (25). HIV can attenuate immunity to other infections. The mechanism may differ between co-infecting pathogens but thought to involve a compromised antibody response because of depletion and dysregulation of CD4 helper T cells (26, 27). ART allows people living with HIV to avoid the worst consequences of HIV infection, which are the result of severe depletion of CD4 T cells (28). However, lack of adherence to ART and development of drug resistance mutations leads to ongoing HIV replication, which, if left to persist for years, results in advanced HIV disease mediated immune suppression (28). We and others have recently shown that advanced HIV disease may lead to delayed clearance of SARS-CoV-2 and evolution of the SARS-CoV-2 virus (29, 30), similar to what is observed in immune suppression caused by other factors (31-35).

Here we mapped neutralization of ancestral, Beta, Alpha and Delta variant viruses and a virus evolved in advanced HIV disease immune suppression by antibodies elicited by each strain or variant. These strains were isolated from infections in South Africa. The SARS-CoV-2 antibody neutralization response tested was from the blood of convalescent individuals infected in one of three SARS-CoV-2 infection waves in South Africa, where the first wave was composed of infections by ancestral strains with the D614G substitution, the second dominated by the Beta variant, and the third by the Delta variant (Fig 1A). We obtained viral isolates from the upper respiratory tract and blood derived plasma from convalescent individuals for each infection wave, and we sequenced viruses eliciting plasma immunity to validate the infecting virus where possible (Table S1). A phylogenetic tree of the variants shows the genetic relationships between the Alpha, Beta, Gamma, Delta and Lambda variants (Fig 1B), as well as the virus evolved from infection of an ancestral SARS-CoV-2 strain in a person with advanced HIV disease (described below).

**Figure 1:**
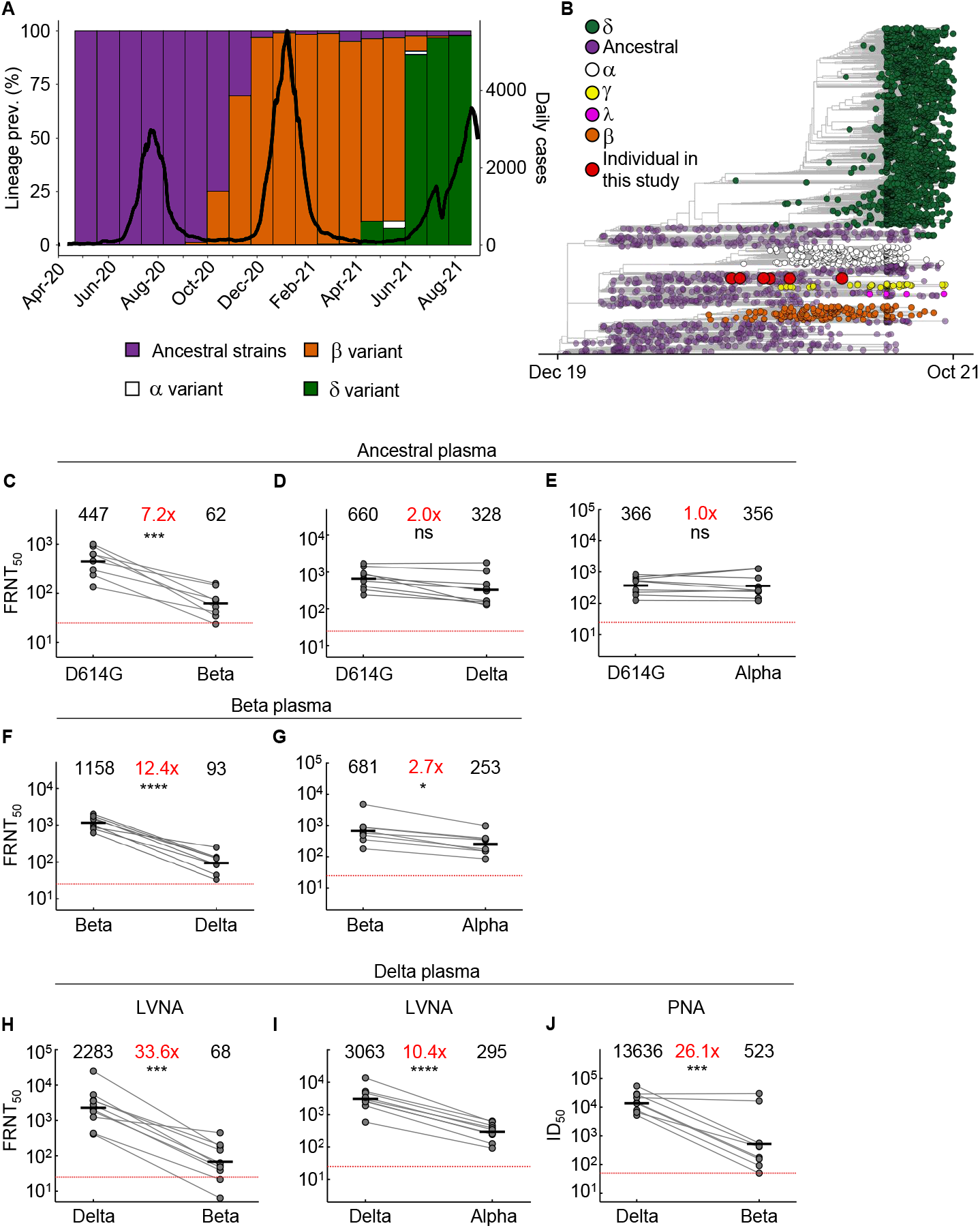
Neutralization distance between variants. (A) Infection waves and variant frequencies in South Africa. (B) Maximum-likelihood phylogenetic tree with evolved virus sequences (red) at 6 time-points in relation to 3883 global sequences with variants shown. (C-E) Neutralization of the Beta (C), Delta (D) and Alpha (E) virus compared to D614G ancestral virus by plasma from convalescent participants infected by ancestral strains (n=8). (F-G) Neutralization of the Delta (F) and Alpha (G) compared to Beta virus by plasma from Beta infections (n=9). (H-I) Neutralization of the Beta (H) and Alpha (I) compared to Delta virus by plasma from Delta infections (n=10). Experiments presented in panels C-I performed by a live virus neutralization assay (LVNA). (J) Neutralization of Beta compared to Delta virus same plasma as (I) using a pseudo-virus neutralization assay (PNA). Red horizontal line denotes most concentrated plasma tested. Numbers in black above each virus strain are geometric mean titers (GMT) of the reciprocal plasma dilution (FRNT50 for LVNA, ID50 for PNA) for 50% neutralization. Numbers in red denote fold-change in GMT between virus strain on the left and the virus strain on the right. p-Values are * <0.05-0.01; ** <0.01-0.001; *** < 0.001-0.0001, **** < 0.0001 as determined by the Wilcoxon rank sum test.

We used a live virus neutralization assay (LVNA) to quantify neutralization. LVNA reads out as the reduction in the number of infection foci at different neutralizing plasma dilutions which is used to obtain the plasma dilution needed for 50% inhibition. We report focus reduction neutralization test (FRNT)50, the reciprocal of this dilution (2). We observed that neutralization capacity of ancestral virus elicited plasma declined 7.2-fold against the Beta virus (Fig 1C). In contrast, it declined only two-fold against the Delta virus (Fig 1D). Neutralization did not decline against the Alpha variant (Fig 1E). Using Beta-elicited plasma, we observed a 12.4-fold decline of neutralization of Delta virus relative to Beta virus (Fig 1F). Alpha virus was well neutralized by Beta elicited plasma, with a 2.7-fold decline compared to the matched Beta virus (Fig 1G). A dramatic decline was observed when Delta plasma was used to neutralize Beta virus. This resulted in a 33.6-fold drop compared to Delta virus neutralization (Fig 1H). Alpha also showed relatively high neutralization escape from Delta elicited plasma, at 10.4-fold relative to Delta virus (Fig 1I), although this was considerably lower than Beta escape. Because we have not previously observed the degree of escape in terms of fold-change seen with Beta virus neutralization by Delta elicited plasma, we repeated the experiments using a pseudovirus neutralization assay (10). We observed increased sensitivity to neutralization with this system relative to LVNA. However, the drop in neutralization of Beta virus with Delta plasma, at 26.1-fold, was very similar to the LVNA results (Fig 1J).

We next characterized SARS-CoV-2 that had evolved in a person with advanced HIV diagnosed in late September 2020 with SARS-CoV-2 infected with the ancestral lineage B.1.1.273 which we previously described in a case report (30). The study participant was discharged following clinical recovery 10 days post-diagnosis according to South Africa guidelines and remained asymptomatic for most study visits. HIV viremia persisted up to day 190 post-diagnosis due to irregular ART adherence and SARS-CoV-2 titer was high, ranging from a Ct of 16 to 27 (30). Diagnosis of SARS-CoV-2 infection was 16 days post-symptom onset and study enrollment was 6 days post-diagnosis (30). Last positive qPCR result was early May 2021. Phylogenetic mapping is consistent with a single infection event (Fig 1B). The CD4 count was <10 at enrollment (Fig 2A top row). It increased at later timepoints, possibly due to the improved adherence to ART and a switch to dolutegravir based therapy which reduced the HIV viral load to below the level of clinical detection (30). SARS-CoV-2 was detected by qPCR until day 216 post-diagnosis (Fig 2A second row). We attempted to isolate live virus up to and including the day 216 post-diagnosis swab sample. While there was insufficient sample to isolate virus from the day 0 swab, we isolated and expanded SARS-CoV-2 from subsequent swabs until and including day 190 post-SARS-CoV-2 diagnosis (Fig 2A third row). Successful isolation indicates that live virus was shed at that time. RBD specific IgG antibodies in the blood were at borderline detection levels (slightly above the mean negative control + 2 std) at the early timepoints but were detected at higher levels starting day 190 (Fig 2A fourth row, Fig S1).

**Figure 2:**
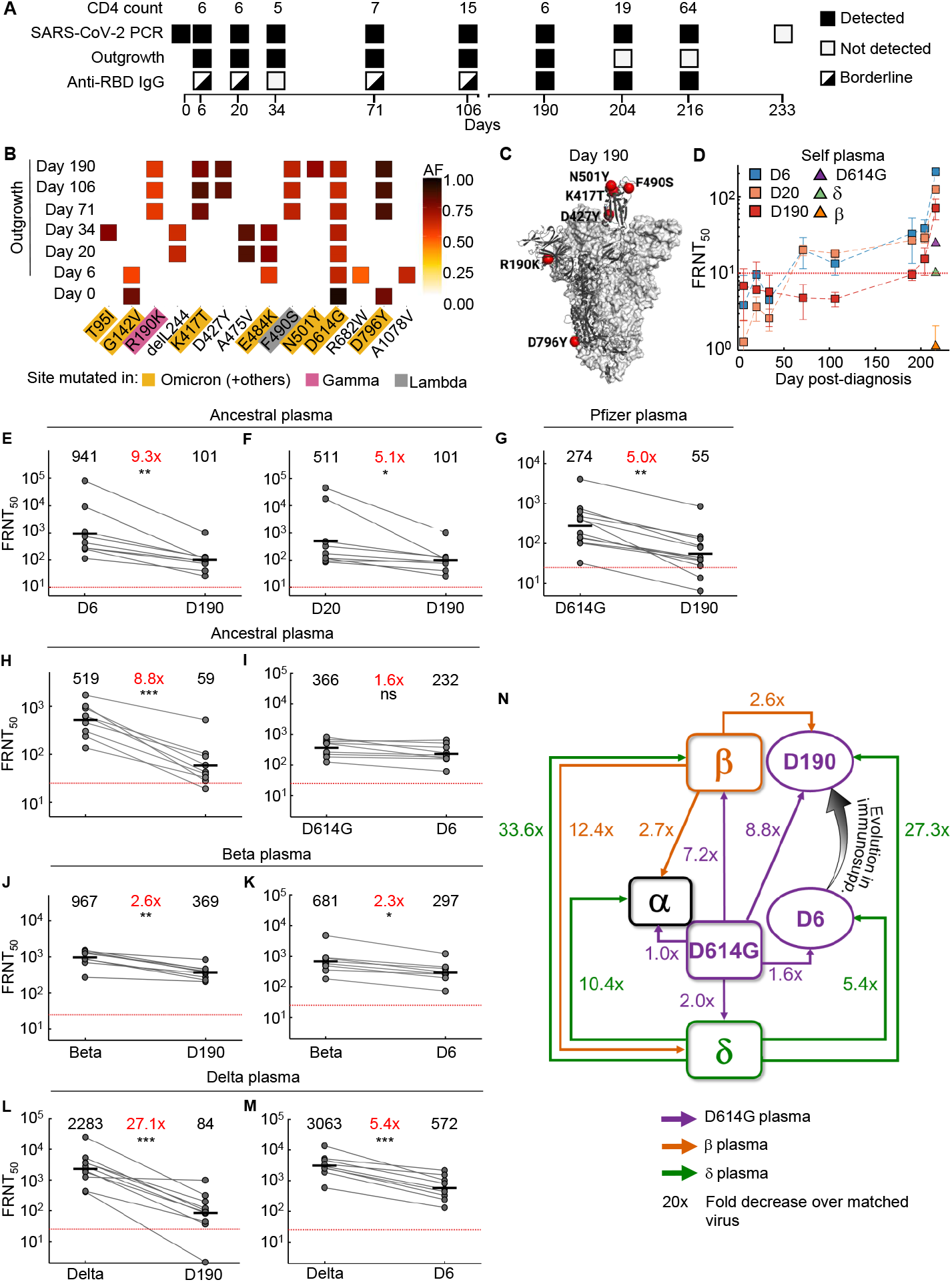
Mapping neutralization of variants and evolved virus. (A) Participant characteristics over 233 days from SARS-CoV-2 diagnosis: CD4 T cell count (cells/µL), SARS-CoV-2 by qPCR, virus outgrowth, and presence of anti-RBD IgG. Because IgG levels were close to the background for some timepoints, they were marked as borderline. (B) Majority and minority SARS-CoV-2 genotypes in the swab (day 0) and outgrowth (day 6 to 190). X-axis lists substitutions and deletions in spike sequence and positions where mutations are found in variants are highlighted. AF: allele frequency. (C) Cryo-EM structure of the SARS-CoV-2 spike protein. The mutations in day 190 isolated virus (D190) shown as red spheres. (D) Neutralization of day 6 isolated (D6), day 20 isolated (D20), and D190 virus by self-plasma collected days 6 to 216 and the ancestral D614G, Beta and Delta viruses with plasma collected day 216. (E-F) Neutralization of D6 (E) and D20 (F) relative to D190 virus by ancestral infection elicited plasma (n=8). (G) Neutralization of D190 compared to D614G by Pfizer BNT162b2 plasma (n=12). (H-I) Neutralization of D190 (H) and D6 (I) compared to D614G by ancestral plasma (n=8). (J-K) Neutralization of D190 (J) and D6 (K) compared to Beta by Beta plasma (n=9). (L-M) Neutralization of D190 (L) and D6 (M) compared to Delta by Delta plasma (n=10). Red horizontal line denotes most concentrated plasma tested. Numbers in black are GMT FRNT50. Numbers in red are fold-change in GMT between virus strain on left and right. p-values are * <0.05-0.01; ** <0.01-0.001, *** <0.001-0.0001 as determined by the Wilcoxon rank sum test. (N) Summary map (not to scale) of serological distances as measured by fold-decrease in neutralization. For clarity, Beta plasma neutralization of D6 not shown.

Outgrown virus was sequenced to detect majority and minority variants (Fig 2B). The mutations found in the outgrown virus were representative of the virus in the swab from the matched time point (Tables S2-S3) except for the R682W substitution at the furin cleavage site on the day 6 sample. This mutation evolves *in vitro* during expansion in VeroE6 cells and likely confers moderate neutralization escape (36). E484K was first detected on day 6 (Fig 2B). This mutation persisted at day 20 and 34 but was replaced with the F490S substitution starting on day 71 when the K417T mutation was also detected. The N501Y mutation was detected in the virus isolated on day 190 post-diagnosis. Mutations were clustered in the RBD, including K417T, F490S, and N501Y in the day 190 viral isolate (Fig 2C, see Fig S2 for mutations per timepoint). Among the RBD mutations in the day 190 isolate, K417T is found in the Gamma variant, and F490S is found in the Lambda variant. Among NTD mutations, T95I is found in Mu, and R190K is at the same location as the R190S in Gamma. N501Y is found in Beta, among others. Interestingly the Omicron variant has emerged as this work was being revised and has mutations at many of the same sites as the evolving virus described here, also shared with some of the other variants (https://covdb.stanford.edu/page/mutation-viewer/#sec_b-1-351). This includes the D796Y mutation only found in Omicron among the major variants (Fig 2B).

We tested three of the isolates for neutralization: viruses outgrown from day 6 and day 20 swabs (designated D6, D20) representing viruses from early infection, and virus outgrown from the day 190 swab (D190), after substantial evolution. Neutralization of the D6, D20, and D190 isolates by self-plasma was low at the early timepoints (Fig 2D). However, neutralization of D6 and D20 was evident by plasma sampled from day 190 and was more pronounced in the plasma sampled from day 216. The D6 isolate was the most sensitive to neutralization by day 216 plasma. Neutralization declined for D20 and further for D190, suggesting sequential evolution of escape (Fig 2D). The ancestral virus and Beta and Delta variants were also tested for neutralization using day 216 plasma. Neutralization was lower for all three non-self strains relative to self-derived virus. The strongest neutralization was of ancestral virus. Delta was neutralized to a lesser degree, and Beta was not detectably neutralized (Fig 2D).

We also tested the D6, D20, and D190 isolates against plasma from other convalescent participants infected with ancestral virus. Neutralization of D190 by ancestral infection elicited plasma was decreased dramatically relative to D6, with FRNT50 for D190 being 9.3-fold lower despite the presence of the E484K mutation in D6 (Figure 2E). The difference was smaller between D190 and D20 (5.1-fold, Figure 2F), consistent with evolution of some neutralization escape in D20 relative to D6. We also tested neutralization of D190 virus using Pfizer BNT162b2 vaccinated participants (Table S4 lists participant characteristics). BNT162b2-elicited plasma neutralization capacity was decreased 5-fold against D190 relative to ancestral virus with the D614G mutation (Fig 2G). We compared neutralization of Beta, D6, D20, and D190 on a subset of remaining BNT162b2 plasma samples from 5 participants 5-6 months post-vaccine, where neutralization declined to relatively low levels. Despite this limitation, neutralization showed a pattern consistent with the other results: D190 neutralization escape was very similar to Beta, while D6 and D20 showed no escape from BNT162b2 elicited neutralization (Fig S3). A 5-fold reduction is less than the fold-drop we previously obtained for the Beta variant with convalescent plasma from previous infection (9) and confirmed below. Moreover, BNT162b2 is effective against Beta, albeit with some reduction. Therefore, these results are consistent with substantial but incomplete escape of D190 from BNT162b2.

We next assessed the serological distance between D190, D6, the ancestral strain, Beta, Delta, and Alpha variants. We tested against convalescent plasma obtained from participants infected with ancestral strains or Beta or Delta variants. Neutralization by ancestral plasma immunity declined 8.8-fold relative to ancestral virus for the D190 isolate (Fig 2H), similar to the Beta variant. In contrast, there was only a 1.6-fold decline for the D6 isolate (Fig 2I). D190 virus was neutralized relatively well by Beta variant infection elicited plasma, with a 2.6-fold reduction (Fig 2J). This was similar to D6 neutralization, which was reduce 2.3-fold relative to Beta (Fig 2K). A much more dramatic decline was observed with Delta variant elicited immunity: a 27.1-fold drop in neutralization capacity compared to neutralization of Delta virus (Fig 2L). Escape from Delta elicited immunity was much more moderate for D6, with a 5.4-fold decline compared to Delta virus (Fig 2M).

Mapping the results (Fig 2N) shows that the Beta and Delta variants are serologically far apart, with ancestral virus forming a hub. The greater distance of Beta relative to Delta from ancestral is consistent with Beta being an escape variant and so evolving antibody escape mutations in the RBD as well as mutations and deletions in the NTD. The Alpha variant was serologically like the ancestral strain and was well neutralized by Beta plasma. However, it did escape Delta elicited neutralization. D6 was serologically similar to the ancestral strain even though it had the E484K substitution, and the in vitro evolved R682W which is reported to confer a moderate decrease in neutralization. In contrast, the D190 virus, which underwent extensive evolution, was serologically like Beta despite sequence differences. It had escape from ancestral and a much more pronounced escape from Delta elicited plasma, similar to the Beta virus. We did not map Gamma, but neutralization escape of the Delta but not the Beta variant from Gamma elicited neutralization (17) may suggest that Beta and Gamma are serologically similar.

We have shown that Delta, Beta, Alpha, and the D190 virus we characterize here, are all escape variants/strains in the sense that each can escape from neutralizing immunity elicited by at least one other variant. This is even though D190 virus and Beta and Alpha variants predate Delta and so Delta escape could not have been selected for. Furthermore, Delta shows weak and Alpha no escape from immunity elicited by ancestral strains, indicating they may have evolved to better transmit and not to escape. This could happen because antibodies are elicited to preferred sites on spike and these differ between variants. Antibodies against the RBD of ancestral strains are concentrated around the E484 site (class 2) which differs between ancestral and Beta (19). In contrast, Beta elicits a stronger response to the class 3 epitope spanning sites 443 to 452 in the RBD (37), where Beta and ancestral do not differ and cross-neutralization by Beta immunity is effective. Delta and Beta do differ at class 3 sites, possibly leading to Delta escape of Beta immunity. Differences in the NTD would further decrease cross-neutralization.

D190 did show escape from BNT162b2 elicited neutralization, although a 5-fold reduction is less than the fold-drop we obtain for the Beta variant with convalescent ancestral plasma. BNT162b2 is effective against Beta, albeit with some reduction (38). Therefore, these results are consistent with substantial but incomplete escape of D190 from BNT162b2.

The driving force behind the evolution of D190 may have been the presence of very low levels of antibody to SARS-CoV-2 which may select antibody escape mutations. While the participant described here did develop a neutralization response and did clear the virus, the response was strongest against the ancestral-like virus early in infection and weak for the Beta-like D190 isolate, indicating a considerable lag between SARS-CoV-2 evolution and neutralization. Consistent with this, neutralization was detectable for the ancestral strain but not Beta virus. Delta virus neutralization was also low. All non-participant derived viruses were neutralized more poorly than participant derived viruses. This may be a source of vulnerability for future SARS-CoV-2 infections in the participant.

A limitation of this study is that we have characterized one case. Out of 93 PLWH in our cohort at the time of analysis (25), 13 had persistent CD4<200 but only one (∼1%) showed extensive evolution of SARS-CoV-2 as described here. However, as there are about 8 million PLWH in South Africa, (https://www.unaids.org/en/regionscountries/countries/southafrica), assuming a frequency of 1% would translate to 80,000 people where SARS-CoV-2 evolution could occur. We have previously reported on a 2-fold decrease in the frequency of detectable HIV antiretroviral therapy and 2-fold increase in detectable HIV viremia in SARS-CoV-2 infected people in the second versus first infection wave in South Africa (25). If immunosuppression by advanced HIV drives SARS-CoV-2 evolution, ART coverage should be increased to prevent it.

Antigenic cartography has been extensively used in influenza (39). Genetic distance may be partially but not completely a proxy for antigenic distance, as some substitutions can lead to large changes in antigenicity while others lead to minor effects (39). It is unclear whether genetic distance is a good measure for antigenic differences between SARS-CoV-2 variants. For example, based on the phylogenetic tree (Fig 1B) it is not obvious that Beta elicited immunity would neutralize Alpha virus better than Delta immunity. Perhaps this is because SARS-CoV-2 evolution seems be closer to HIV, where multiple strains radiate from a common ancestor (40), than to the stepwise progression in influenza (39). How far the variants will diverge for each other is unclear, but if further divergence occurs, SARS-CoV-2 may form serotypes such as occur with polio (41) and dengue (42) viruses. With serotypes, antibody immunity is developed to the infecting but not the other serotypes. This may imply that a vaccine which is based on one variant or strain or previous infection with that variant may generate immunity which is vulnerable to infection by another variant.

What surprised us in the participant with prolonged infection was that, over the 6-month period where SARS-CoV-2 titer was high, infection was for the most part asymptomatic. One explanation is that the virus evolved attenuated pathogenicity or had low pathogenicity to begin with. Another may that high pathogen levels over time are tolerated because immunosuppression reduces the inflammatory response. This is observed in tuberculosis (43). No attenuated pathogenicity was observed with the Beta escape variant (25).

As this work is being revised, the Omicron variant has been detected in South Africa. Omicron (B.1.1.529) is derived from strains which were common in the first infection wave in South Africa, 1.5 years ago, but were then supplanted first by Beta, then Delta. There may be several possibilities for how this virus persisted so long without being detected. One explanation is that Omicron may have evolved in a region with less developed genomic surveillance relative to South Africa and may have arrived in South Africa recently. Alternatively and without excluding the former possibility, it may have persisted in a single immune compromised host until it acquired a critical mass of mutations that would allow it to effectively transmit in a population where the prevalence of previous infection is high (44, 45). In support of this, previous studies investigating SARS-CoV-2 evolution in people immune-compromised for reasons other than advanced HIV disease have established that such evolution leads to immune escape mutations (31, 33). Therefore, evolution of escape is not related directly to HIV but rather to advanced unsuppressed disease which leads to severe damage to the immune response. If the B.1.1.273 strain described in this work and Omicron share a common evolutionary mechanism, Omicron may have Beta-like serology, substantial but incomplete escape from neutralizing immunity elicited by mRNA vaccines, and strong escape of Delta elicited immunity which may lead to reinfections. Whether other features could also be shared, such as potentially mild pathogenicity, is yet unknown.

## Methods

### Ethical statement

Nasopharyngeal and oropharyngeal swab samples and blood samples were obtained from hospitalized adults with PCR-confirmed SARS-CoV-2 infection or vaccinated individuals who were enrolled in a prospective cohort study approved by the Biomedical Research Ethics Committee at the University of KwaZulu–Natal (reference BREC/00001275/2020).

### Whole-genome sequencing, genome assembly and phylogenetic analysis

cDNA synthesis was performed on the extracted RNA using random primers followed by gene-specific multiplex PCR using the ARTIC V.3 protocol (https://www.protocols.io/view/covid-19-artic-v3-illumina-library-construction-an-bibtkann). In brief, extracted RNA was converted to cDNA using the Superscript IV First Strand synthesis system (Life Technologies) and random hexamer primers. SARS-CoV-2 whole-genome amplification was performed by multiplex PCR using primers designed using Primal Scheme (http://primal.zibraproject.org/) to generate 400-bp amplicons with an overlap of 70 bp that covers the 30 kb SARS-CoV-2 genome. PCR products were cleaned up using AmpureXP purification beads (Beckman Coulter) and quantified using the Qubit dsDNA High Sensitivity assay on the Qubit 4.0 instrument (Life Technologies). We then used the Illumina Nextera Flex DNA Library Prep kit according to the manufacturer’s protocol to prepare indexed paired-end libraries of genomic DNA. Sequencing libraries were normalized to 4 nM, pooled and denatured with 0.2 N sodium acetate. Then, a 12-pM sample library was spiked with 1% PhiX (a PhiX Control v.3 adaptor-ligated library was used as a control). We sequenced libraries on a 500-cycle v.2 MiSeq Reagent Kit on the Illumina MiSeq instrument (Illumina). We assembled paired-end fastq reads using Genome Detective 1.126 (https://www.genomedetective.com) and the Coronavirus Typing Tool. We polished the initial assembly obtained from Genome Detective by aligning mapped reads to the reference sequences and filtering out low-quality mutations using the bcftools 1.7-2 mpileup method. Mutations were confirmed visually with BAM files using Geneious software (Biomatters). We analyzed sequences from the six different time points (D6, D20, D34, D71, D106 and D190) against a global reference dataset of 3883 genomes using a custom build of the SARS-CoV-2 NextStrain (https://github.com/nextstrain/ncov). The workflow performs alignment of genomes, phylogenetic tree inference, tree dating and ancestral state construction and annotation. The phylogenetic tree was visualized using ggplot and ggtree. All sequences from the participant clustered in a monophyletic clade (https://nextstrain.org/groups/ngs-sa/COVID19-AHRI-2021.05.27?label=clade:HIV%20patient) that are well separated from the rest of the phylogeny.

### Cells

Vero E6 cells (ATCC CRL-1586, obtained from Cellonex in South Africa) were propagated in complete DMEM with 10% fetal bovine serum (Hylone) containing 1% each of HEPES, sodium pyruvate, L-glutamine and nonessential amino acids (Sigma-Aldrich). Vero E6 cells were passaged every 3–4 days. The H1299-E3 cell line for first-passage SARS-CoV-2 expansion, derived as described in (9), was propagated in complete RPMI with 10% fetal bovine serum containing 1% each of HEPES, sodium pyruvate, L-glutamine and nonessential amino acids. H1299 cells were passaged every second day. Cell lines have not been authenticated. The cell lines have been tested for mycoplasma contamination and are mycoplasma negative.

### Virus expansion

All work with live virus was performed in Biosafety Level 3 containment using protocols for SARS-CoV-2 approved by the AHRI Biosafety Committee. We used ACE2-expressing H1299-E3 cells for the initial isolation (P1 stock) followed by passaging in Vero E6 cells (P2 and P3 stocks, where P3 stock was used in experiments). ACE2-expressing H1299-E3 cells were seeded at 4.5 × 10^5^ cells in a 6 well plate well and incubated for 18–20 h. After one DPBS wash, the sub-confluent cell monolayer was inoculated with 500 μL universal transport medium diluted 1:1 with growth medium filtered through a 0.45-μm filter. Cells were incubated for 1 h. Wells were then filled with 3 mL complete growth medium. After 8 days of infection, cells were trypsinized, centrifuged at 300 rcf for 3 min and resuspended in 4 mL growth medium. Then 1 mL was added to Vero E6 cells that had been seeded at 2 × 10^5^ cells per mL 18–20 h earlier in a T25 flask (approximately 1:8 donor-to-target cell dilution ratio) for cell-to-cell infection. The coculture of ACE2-expressing H1299-E3 and Vero E6 cells was incubated for 1 h and the flask was then filled with 7 mL of complete growth medium and incubated for 6 days. The viral supernatant (P2 stock) was aliquoted and stored at −80 °C and further passaged in Vero E6 cells to obtain the P3 stock used in experiments.

### Live virus neutralization assay

Vero E6 cells were plated in a 96-well plate (Corning) at 30,000 cells per well 1 day pre-infection. Plasma was separated from EDTA-anticoagulated blood by centrifugation at 500 rcf for 10 min and stored at −80 °C. Aliquots of plasma samples were heat-inactivated at 56 °C for 30 min and clarified by centrifugation at 10,000 rcf for 5 min. GenScript A02051 anti-spike neutralizing monoclonal antibody was added as a positive control to one column of wells. Virus stocks were used at approximately 50-100 focus-forming units per microwell and added to diluted plasma. Antibody–virus mixtures were incubated for 1 h at 37 °C, 5% CO_2_. Cells were infected with 100 μL of the virus–antibody mixtures for 1 h, then 100 μL of a 1X RPMI 1640 (Sigma-Aldrich, R6504), 1.5% carboxymethylcellulose (Sigma-Aldrich, C4888) overlay was added without removing the inoculum. Cells were fixed 18 h post-infection using 4% PFA (Sigma-Aldrich) for 20 min. Foci were stained with a rabbit anti-spike monoclonal antibody (BS-R2B12, GenScript A02058) at 0.5 μg/mL in a permeabilization buffer containing 0.1% saponin (Sigma-Aldrich), 0.1% BSA (Sigma-Aldrich) and 0.05% Tween-20 (Sigma-Aldrich) in PBS. Plates were incubated with primary antibody overnight at 4 °C, then washed with wash buffer containing 0.05% Tween-20 in PBS. Secondary goat anti-rabbit horseradish peroxidase (Abcam ab205718) antibody was added at 1 μg/mL and incubated for 2 h at room temperature with shaking. TrueBlue peroxidase substrate (SeraCare 5510-0030) was then added at 50 μL per well and incubated for 20 min at room temperature. Plates were imaged in an ELISPOT instrument with built-in image analysis (C.T.L).

### Pseudovirus neutralization assay

SARS-CoV-2 pseudotyped lentiviruses were prepared by co-transfecting the HEK 293T cell line with either the SARS-CoV-2 Beta spike (L18F, D80A, D215G, K417N, E484K, N501Y, D614G, A701V, 242-244 del) or the Delta spike (T19R, R158G L452R, T478K, D614G, P681R, D950N, 156-157 del) plasmids in conjunction with a firefly luciferase encoding lentivirus backbone plasmid. For the neutralization assay, heat-inactivated plasma samples from vaccine recipients were incubated with the SARS-CoV-2 pseudotyped virus for 1 hour at 37°C, 5% CO_2_. Subsequently, 1×10^4^ HEK 293T cells engineered to over-express ACE-2 were added and incubated at 37°C, 5% CO_2_ for 72 hours upon which the luminescence of the luciferase gene was measured. CB6 was used as a positive control.

### Receptor binding domain ELISA

Plasma samples were tested for anti-SARS-CoV-2 IgG. Flat bottom microplates (ThermoFisher Scientific) were coated with 500 ng/mL of the receptor binding domain (RBD) protein (provided by Dr Galit Alter from the Ragon Institute) and incubated overnight at 4°C. Plates were blocked with a 200 µL/well tris-buffered saline containing 1% BSA (TBSA) and incubated at room temperature (RT) for 1h. Samples were diluted in TBSA with 0.05% Tween-20 (TBSAT) to 1:100. Subsequently, goat anti-human IgG (1:5000)-horseradish peroxidase conjugated secondary antibodies (Jackson ImmunoResearch) were added a 100 µL/well and incubated at RT for 1h. Bound secondary antibodies were detected using 100 μL/well 1-step Ultra TMB substrate (ThermoFisher Scientific). Plates were incubated at RT for 3 min in the dark before addition of 1 N sulphuric acid stop solution at 100 μL/well. Plates were washed with 1X high salt TBS containing 0.05% Tween-20, three times each after coating and blocking, and five times each after the sample and secondary antibody. The concentration of anti-RBD expressed as ng/mL equivalent of anti-SARS-CoV-2 monoclonal, CR3022 (Genscript). We used pre-pandemic plasma samples as negative controls to define seroconversion cut-offs calculated as mean + 2 std of the negative samples.

### Statistics and fitting

All statistics and fitting were performed using MATLAB v.2019b. Neutralization data were fit to Tx=1/1+(D/ID50).

Here Tx is the number of foci normalized to the number of foci in the absence of plasma on the same plate at dilution D and ID50 is the plasma dilution giving 50% neutralization. FRNT50 = 1/ID50. Values of FRNT50 <1 are set to 1 (undiluted), the lowest measurable value.

## Data Availability

All data available in the manuscript, GISAID SARS-CoV-2 sequence repository, or upon reasonable request from the authors.

https://www.gisaid.org/

## Acknowledgements

This study was supported by the Bill and Melinda Gates award INV-018944 (AS), National Institutes of Health award R01 AI138546 (AS), South African Medical Research Council awards (AS, TdO, PLM) and National Institutes of Health U01 AI151698 (WVV). PLM is supported by the South African Research Chairs Initiative of the Department of Science and Innovation and the NRF (Grant No 9834). The funders had no role in study design, data collection and analysis, decision to publish, or preparation of the manuscript. We thank Hylton Rodel for advice on figure preparation.

## COMMIT-KZN Team

Moherndran Archary, Department of Paediatrics and Child Health, University of KwaZulu-Natal, Durban, South Africa.

Philip Goulder, Africa Health Research Institute and Department of Paediatrics, Oxford, UK.

Nokwanda Gumede, Africa Health Research Institute, Durban, South Africa.

Ravindra K. Gupta, Africa Health Research Institute and Cambridge Institute of Therapeutic Immunology & Infectious Disease, Cambridge, UK.

Guy Harling, Africa Health Research Institute and the Institute for Global Health, University College London, London, UK.

Rohen Harrichandparsad, Department of Neurosurgery, University of KwaZulu-Natal, Durban, South Africa.

Kobus Herbst, Africa Health Research Institute and the South African Population Research Infrastructure Network, Durban, South Africa.

Prakash Jeena, Department of Paediatrics and Child Health, University of KwaZulu-Natal, Durban, South Africa.

Zesuliwe Jule, Africa Health Research Institute, Durban, South Africa.

Thandeka Khoza, Africa Health Research Institute, Durban, South Africa.

Nigel Klein, Africa Health Research Institute and the Institute of Child Health, University College London, London, UK.

Henrik Kloverpris, Africa Health Research Institute, Durban, South Africa.

Alasdair Leslie, Africa Health Research Institute, Durban, South Africa.

Rajhmun Madansein, Department of Cardiothoracic Surgery, University of KwaZulu-Natal, Durban, South Africa.

Mohlopheni Marakalala, Africa Health Research Institute, Durban, South Africa.

Yoliswa Miya, Africa Health Research Institute, Durban, South Africa.

Mosa Moshabela, College of Health Sciences, University of KwaZulu-Natal, Durban, South Africa.

Nokukhanya Msomi, Department of Virology, University of KwaZulu-Natal, Durban, South Africa.

Kogie Naidoo, Centre for the AIDS Programme of Research in South Africa, Durban, South Africa.

Zaza Ndhlovu, Africa Health Research Institute, Durban, South Africa.

Kennedy Nyamande, Department of Pulmonology and Critical Care, University of KwaZulu-Natal, Durban, South Africa.

Vinod Patel, Department of Neurology, University of KwaZulu-Natal, Durban, South Africa.

Dirhona Ramjit, Africa Health Research Institute, Durban, South Africa.

Kajal Reedoy, Africa Health Research Institute, Durban, South Africa.

Theresa Smit, Africa Health Research Institute, Durban, South Africa.

Adrie Steyn, Africa Health Research Institute, Durban, South Africa.

Emily Wong, Africa Health Research Institute, Durban, South Africa.

## Supplementary materials

**Fig S 1:**
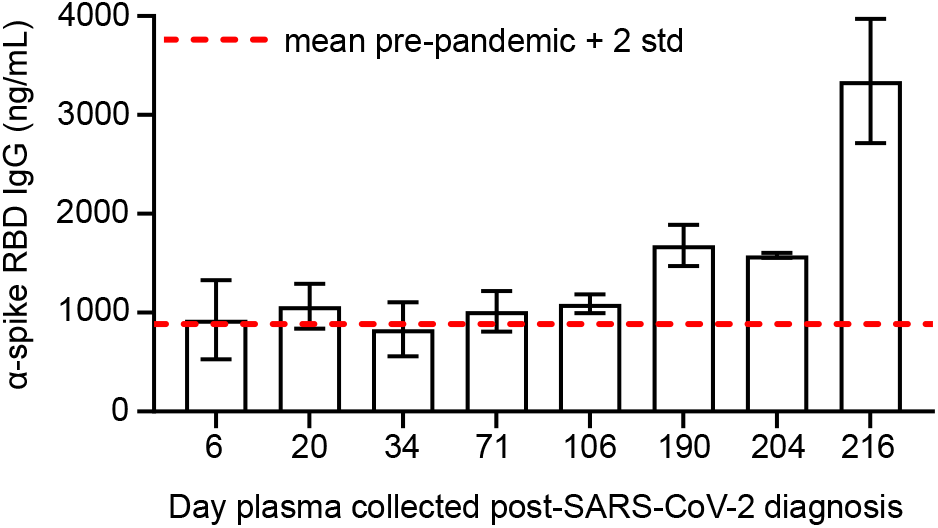
Spike specific antibody levels with time post-SARS-CoV-2 diagnosis. Shown are mean (n=4 replicates per timepoint) and standard deviation of anti-spike RBD antibody concentrations measured in the plasma of the participant with advanced HIV disease by ELISA. Red dashed line denotes the mean + 2 standard deviations of signal from a set of 6 control samples, including plasma of pre-pandemic controls (n=4) and pre-pandemic commercial human serum (n=2).

**Fig S 2:**
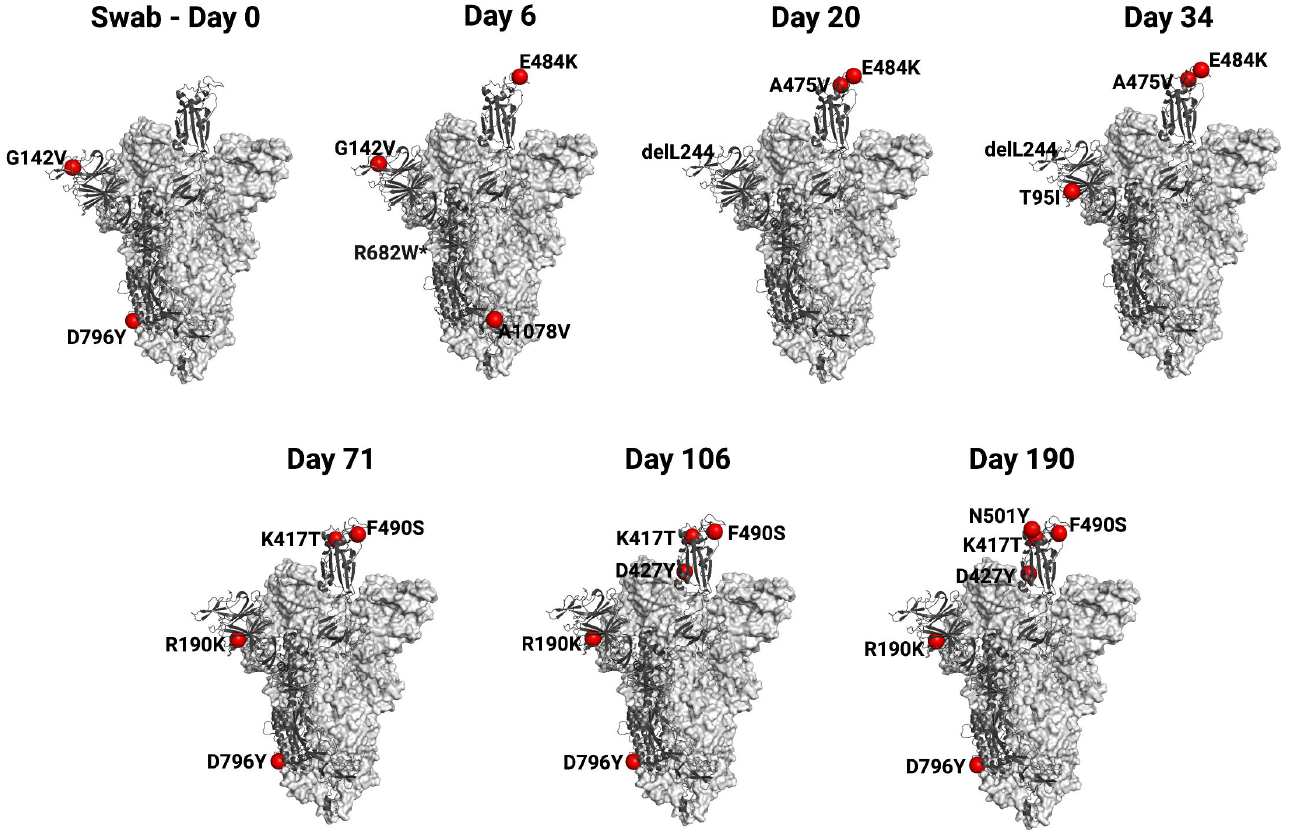
Location of spike substitutions at different timepoints. Cryo-EM structure of the SARS-CoV-2 spike protein with one RBD protomer in the “up” conformation (PDB ID: 7A94). The mutations which accumulated in the SARS-CoV-2-infected immunocompromised individual over 190 days are shown in red spheres. One protomer is shown in cartoon representation (dark grey) and the other two protomers in surface representation (light grey). Analysis of the structure was performed in Pymol version 2.0.7. The R682 residue is not present in the structure.

**Fig S 3:**
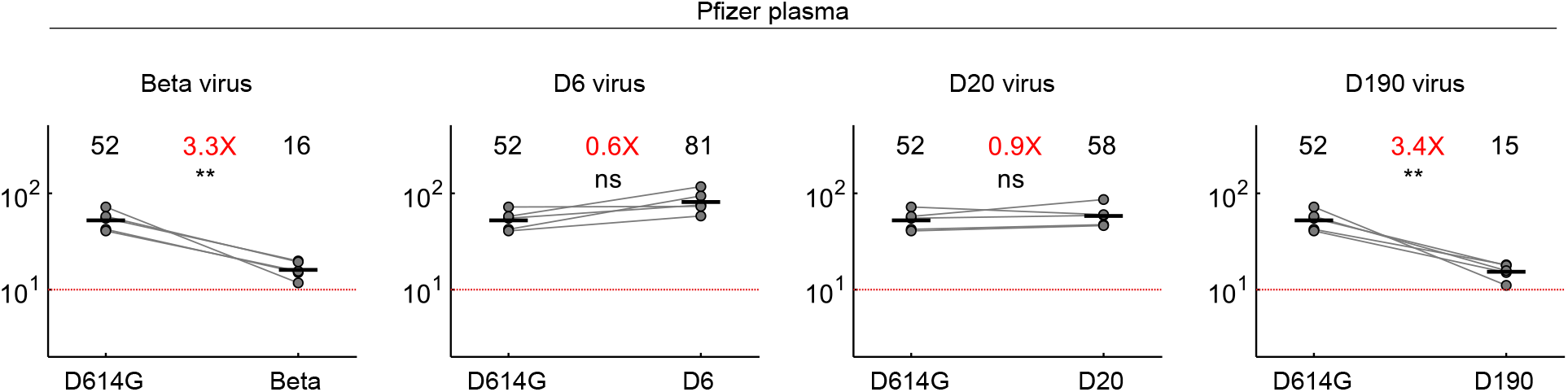
Neutralization of Beta variant and evolved virus by a subset of BNT162b2 plasma donors. Neutralization of Beta, D6, D20, and D190 compared to D614G by Pfizer BNT162b2 plasma (n=5). Plasma donors were 136072, 136074, 136075, 136076, 136078. Red horizontal line denotes most concentrated plasma tested. Numbers in black are GMT FRNT50. Numbers in red are fold-change in GMT between virus strain on left and right. ** *<*0.01-0.001, as determined by the Wilcoxon rank sum test.

**Table S1:** Characteristics of SARS-CoV-2 convalescent study participants.

| Cohort ID | Variant/strain by infection date | Variant/strain by sequencing | Sequence ID | GISAID Accession | Age range (y) | Sex | Days between symptom onset and plasma collection |
| --- | --- | --- | --- | --- | --- | --- | --- |
| 039-02-0027 | Ancestral | B.1.1.273 | K008646 | EPI_ISL_2397308 | 30-39 | F | 6, 20, 26, 34, 71, 106, 190 |
| 039-02-0005 | Ancestral | B.1.1 | K003667 | EPI_ISL_602623 | 50-59 | M | 29 |
| 039-02-0011 | Ancestral | B.1.1.273 | K003675 | EPI_ISL_602631 | 40-49 | F | 32 |
| 039-02-0013 | Ancestral | B.1.1.117 | K003668 | EPI_ISL_602624 | 70+ | F | 29 |
| 039-02-0014 | Ancestral | B.1.1 | K004289 | EPI_ISL_660170 | 60-69 | F | 27 |
| 039-02-0017 | Ancestral | B.1.140 | K004295 | EPI_ISL_660176 | 60-69 | F | 28 |
| 039-13-0018 | Ancestral | B.1.1.84 | K003673 | EPI_ISL_602629 | 40-49 | F | 28 |
| 039-13-0033 | Ancestral | B.1 | K004291 | EPI_ISL_660172 | 30-39 | F | 30 |
| 039-13-0062 | Ancestral | C.9 | K004302 | EPI_ISL_660181 | 60-69 | M | 26 |
| 039-02-0030 | Beta | Beta | K008635 | N/A* | 40-49 | F | 30 |
| 039-02-0031 | Beta | Beta | K008636 | N/A* | 40-49 | F | 41 |
| 039-02-0032 | Beta | Beta | K008628 | N/A* | 40-49 | M | 32 |
| 039-02-0033 | Beta | Beta | K008637 | EPI_ISL_1229368 | 50-59 | M | 42 |
| 039-02-0034 | Beta | N/A<sup>§</sup> | N/A<sup>§</sup> | N/A | 30-39 | F | 32 |
| 039-02-0046 | Beta | Beta | K010372 | N/A* | 30-39 | M | 33 |
| 039-02-1011 | Beta | Beta | K010356 | N/A* | 70+ | F | 48 |
| 039-09-0001 | Beta | Beta | K008633 | EPI_ISL_1229367 | 60-69 | F | 29 |
| 039-02-0045 | Beta | Beta | K010370 | N/A* | 30-39 | F | 31 |
| 039-02-0104 | Delta | Delta | K021407 | EPI_ISL_3722338 | 40-49 | F | 26 |
| 039-02-0106 | Delta | Delta | K021399 | EPI_ISL_3722335 | 40-49 | M | 23<sup>#</sup> |
| 039-02-0108 | Delta | Delta | K021401 | N/A* | 50-59 | M | 31 |
| 039-02-0109 | Delta | Delta | K021225 | N/A* | 40-49 | M | 13<sup>#</sup> |
| 039-13-0153 | Delta | Delta | K021400 | N/A* | 40-49 | M | 44 |
| 039-13-0140 | Delta | Delta | K021226 | N/A* | 50-59 | M | 44 |
| 039-13-0141 | Delta | Delta | K020186 | EPI_ISL_3939068 | 40-49 | M | 31 |
| 039-13-0142 | Delta | Delta | K020187 | EPI_ISL_3939088 | 30-39 | M | 31 |
| 039-13-0149 | Delta | Delta | K020214 | EPI_ISL_3447779 | 50-59 | F | 30<sup>#</sup> |
| 039-13-0157 | Delta | Delta | K021404 | N/A* | 30-39 | M | 32 |

\* <90% coverage. Not submitted to GISAID but sufficient sequence for variant call. <sup>§</sup> Not sequenced. <sup>#</sup> Asymptomatic at diagnosis, date post-diagnostic test used instead of symptom onset date.

**Table S2:** Read numbers at nucleotides which led to amino acid substitutions or deletions in virus sequenced from the swab.

| **Amino Acid change** | **Nucleotide change** | **Codon change** | **Day 0** | **Day 6** | **Day 20** | **Day 34** | **Day 71** | **Day 106** | **Day 190** |
| --- | --- | --- | --- | --- | --- | --- | --- | --- | --- |
| <u>T95I</u> | 21846C>T | 21845<br>ACT>ATT | ACT – 143<br>ATT – 0 | ACT – 6105<br>ATT - 9 | ACT – 2667<br>ATT - 298 | ACT – 935<br>ATT – 1626 | ACT – 1379<br>ATT – 36 | ACT –11843<br>ATT – 576 | ACT – 231<br>ATT – 10 |
| <u>G142V</u> | 21987G>T | 21986<br>GGT>GTT | GGT – 10<br>GTT – 40 | GGT – 1386<br>GTT - 622 | GGT – 880<br>GTT - 313 | GGT – 954<br>GTT – 96 | GGT – 480<br>GTT – 5 | GGT – 3621<br>GTT – 12 | GGT – 133<br>GTT – 22 |
| <u>R190K</u> | 22131G>A | 22130<br>AGG>AAG | AGG – 46<br>AAG – 0 | AGG – 5591<br>AAG – 8 | AGG – 1702<br>AAG – 1 | AGG –<br>1685<br>AAG - 2 | AGG – 222<br>AAG – 1096 | Gap in<br>sequence | AGG – 13<br>AAG – 40 |
| <u>*L244del</u> | 22293ACA<br>>del | 22811<br>ACA>del | ACA – 12<br>del – 1 | ACA – 1378<br>del - 0 | ACA – 50<br>del – 158 | ACA – 17<br>del – 176 | ACA – 414<br>del – 4 | ACA – 11<br>del – 0 | ACA – 64<br>del – 0 |
| <u>K417T</u> | 22812A>G | 22811<br>AAG>ACG | AAG – 117<br>ACG – 0 | AAG – 3237<br>ACG - 1 | AAG – 3209<br>ACG – 0 | AAG – 2587<br>ACG – 1 | AAG – 299<br>ACG – 1362 | AAG – 32<br>ACG – 2 | Gap in<br>sequence |
| <u>D427Y</u> | 22841G>T | 22925<br>GAT>TAT | GAT – 107<br>TAT – 0 | GAT – 3856<br>TAT – 3 | GAT – 3719<br>TAT – 0 | GAT – 3434<br>TAT – 0 | GAT – 3434<br>TAT – 0 | GAT – 51<br>TAT – 4 | GAT – 18<br>TAT - 103 |
| <u>L455F</u> | 22927G>T | 22925<br>TTG>TTT | TTG – 11<br>TTT – 0 | TTG – 3714<br>TTT – 9 | TTG – 3219<br>TTT – 0 | TTG – 4331<br>TTT – 2 | TTG –2983<br>TTT – 4 | TTG –527<br>TTT – 317 | TTG – 114<br>TTT – 0 |
| <u>F456L</u> | 22928T>C | 22928<br>TTT>CTT | TTT – 12<br>CTT – 0 | TTT – 3739<br>CTT – 6 | TTT – 3248<br>CTT – 0 | TTT – 4361<br>CTT – 0 | TTT – 2971<br>CTT – 3 | TTT – 537<br>CTT – 310 | TTT – 138<br>CTT – 0 |
| <u>A475V</u> | 22986C>T | 22985<br>GCC>GTC | GCC – 17<br>GTC – 0 | GCC – 3409<br>GTC - 11 | GCC – 3629<br>GTC – 1218 | GCC – 2874<br>GTC – 2421 | GCC – 3396<br>GTC – 101 | GCC – 901<br>GTC – 23 | GCC – 177<br>GTC – 14 |
| <u>E484K</u> | 23012G>A | 23012<br>GAA>AAA | GAA – 17<br>AAA – 0 | GAA – 56<br>AAA - 2063 | GAA – 1632<br>AAA – 2881 | GAA – 2395<br>AAA – 2139 | GAA – 3147<br>AAA – 196 | GAA – 688<br>AAA – 40 | GAA – 180<br>AAA – 53 |
| <u>F490S</u> | 23031T>C | 23030<br>TTT>TCT | TTT – 18<br>TCT – 0 | TTT- 1605<br>TCT - 2 | TTT – 4067<br>TCT – 0 | TTT – 4007<br>TCT – 8 | TTT – 341<br>TCT – 2912 | TTT – 182<br>TCT – 446 | TTT – 89<br>TCT – 125 |
| <u>N501Y</u> | 23063A>T | 23063<br>AAT>TAT | AAT – 13<br>TAT – 0 | AAT – 810<br>TAT – 2 | AAT – 2034<br>TAT – 293 | AAT – 2855<br>TAT – 29 | AAT – 2510<br>TAT – 14 | AAT – 282<br>TAT – 13 | AAT – 19<br>TAT – 180 |
| <u>D614G</u> | 23403A>G | 23402<br>GAT>GGT | GAT – 3<br>GGT – 158 | GAT – 8<br>GGT 6668 | GAT – 0<br>GGT – 5299 | GAT – 3<br>GGT – 2863 | GAT – 3<br>GGT – 1560 | GAT – 1<br>GGT – 26 | GAT – 2<br>GGT – 173 |
| <u>R682W</u> | 23606C>T | 23606<br>CGG>TGG | CGG – 98<br>TGG – 1 | CGG – 1831<br>TGG – 4 | CGG – 27<br>TGG – 0 | CGG – 59<br>TGG – 0 | CGG – 124<br>TGG – 0 | CGG – 12<br>TGG – 1 | CGG – 40<br>TGG – 4 |
| <u>D796Y</u> | 23948G>T | 23948<br>GAT>TAT | GAT – 16<br>TAT – 70 | GAT – 4293<br>TAT – 16 | GAT – 1235<br>TAT – 477 | GAT – 568<br>TAT – 649 | GAT – 97<br>TAT – 791 | GAT – 7<br>TAT – 0 | GAT – 2<br>TAT – 11 |
| <u>A1078V</u> | 24795C>T | 24974<br>GCT>GTT | GCT – 219<br>GTT – 3 | GCT – 772<br>GTT - 4833 | GCT – 4397<br>GTT – 1 | GCT – 2519<br>GTT – 12 | GCT – 1462<br>GTT – 30 | GCT –13695<br>GTT – 74 | GCT – 286<br>GTT – 17 |

\*Only deletions where the adjacent codon was preserved were counted.

**Table S3:** Read numbers at nucleotides which led to amino acid substitutions or deletions in virus sequenced from outgrown stock.

| **Amino Acid change** | **Nucleotide change** | **Codon change** | **Day 0 (Swab)** | **Day 6** | **Day 20** | **Day 34** | **Day 71** | **Day 106** | **Day 190** |
| --- | --- | --- | --- | --- | --- | --- | --- | --- | --- |
| <u>T95I</u> | 21846C>T | 21845<br>ACT>ATT | ACT – 143<br>ATT – 0 | ACT – 1342<br>ATT - 10 | ACT – 1540<br>ATT - 33 | ACT – 96<br>ATT – 1328 | ACT – 1609<br>ATT – 27 | ACT – 1593<br>ATT – 29 | ACT – 1325<br>ATT – 16 |
| <u>G142V</u> | 21987G>T | 21986<br>GGT>GTT | GGT – 10<br>GTT – 40 | GGT – 316<br>GTT - 808 | GGT – 1352<br>GTT - 4 | GGT – 1238<br>GTT – 14 | GGT – 1416<br>GTT – 2 | GGT – 1400<br>GTT – 5 | GGT – 977<br>GTT – 6 |
| <u>R190K</u> | 22131G>A | 22130<br>AGG>AAG | AGG – 46<br>AAG – 0 | AGG – 119<br>AAG – 0 | AGG – 357<br>AAG – 3 | AGG – 324<br>AAG - 2 | AGG – 26<br>AAG – 378 | AGG – 22<br>AAG – 468 | AGG – 419<br>AAG – 33 |
| <u>*L244del</u> | 22293ACA<br>>del | 22811<br>ACA>del | ACA – 12<br>del – 1 | ACA – 371<br>del - 4 | ACA – 0<br>del – 635 | ACA – 17<br>del – 556 | ACA – 692<br>del – 3 | ACA – 693<br>del – 7 | ACA – 653<br>del – 1 |
| <u>K417T</u> | 22812A>G | 22811<br>AAG>ACG | AAG – 117<br>ACG – 0 | AAG – 141<br>ACG - 0 | AAG – 122<br>ACG – 0 | AAG – 96<br>ACG – 0 | AAG – 6<br>ACG – 172 | AAG – 0<br>ACG – 60 | AAG – 0<br>ACG – 17 |
| <u>D427Y</u> | 22841G>T | 22841<br>GAT>TAT | GAT – 107<br>TAT – 0 | GAT – 1281<br>TAT – 7 | GAT – 1273<br>TAT – 1 | GAT – 1164<br>TAT - 1 | GAT – 1079<br>TAT – 42 | GAT – 13<br>TAT – 743 | GAT – 6<br>TAT - 529 |
| <u>L455F</u> | 22927G>T | 22925<br>TTG>TTT | TTG – 11<br>TTT – 0 | TTG – 886<br>TTT – 0 | TTG – 978<br>TTT – 2 | TTG – 888<br>TTT – 3 | TTG – 784<br>TTT – 2 | TTG – 636<br>TTT – 0 | TTG – 454<br>TTT – 1 |
| <u>F456L</u> | 22928T>C | 22928<br>TTT>CTT | TTT – 12<br>CTT – 0 | TTT – 1112<br>CTT – 31 | TTT – 1109<br>CTT – 19 | TTT – 1011<br>CTT – 20 | TTT – 882<br>CTT – 19 | TTT – 725<br>CTT – 2 | TTT – 544<br>CTT – 24 |
| <u>A475V</u> | 22986C>T | 22985<br>GCC>GTC | GCC – 17<br>GTC – 0 | GCC – 1180<br>GTC - 30 | GCC – 11<br>GTC – 1369 | GCC – 43<br>GTC – 1224 | GCC – 956<br>GTC – 22 | GCC – 839<br>GTC – 24 | GCC – 687<br>GTC – 19 |
| <u>E484K</u> | 23012G>A | 23012<br>GAA>AAA | GAA – 17<br>AAA – 0 | GAA – 363<br>AAA - 917 | GAA – 27<br>AAA – 1276 | GAA – 55<br>AAA – 1133 | GAA – 1120<br>AAA – 22 | GAA – 1001<br>AAA – 5 | GAA – 814<br>AAA – 4 |
| <u>F490S</u> | 23031T>C | 23030<br>TTT>TCT | TTT – 18<br>TCT – 0 | TTT- 1196<br>TCT - 15 | TTT – 1226<br>TCT – 10 | TTT – 1031<br>TCT – 30 | TTT – 62<br>TCT – 935 | TTT – 31<br>TCT – 784 | TTT – 21<br>TCT – 669 |
| <u>N501Y</u> | 23063A>T | 23063<br>AAT>TAT | AAT – 13<br>TAT – 0 | AAT – 1428<br>TAT – 2 | AAT – 1498<br>TAT – 1 | AAT – 1354<br>TAT – 0 | AAT – 1193<br>TAT – 2 | AAT – 1026<br>TAT – 0 | AAT – 6<br>TAT – 726 |
| <u>D614G</u> | 23403A>G | 23402<br>GAT>GGT | GAT – 3<br>GGT – 158 | GAT – 4<br>GGT 811 | GAT – 6<br>GGT – 904 | GAT – 14<br>GGT – 886 | GAT – 6<br>GGT – 702 | GAT – 10<br>GGT – 827 | GAT – 3<br>GGT – 834 |
| <u>R682W</u> | 23606C>T | 23606<br>CGG>TGG | CGG – 98<br>TGG – 1 | CGG – 189<br>TGG – 407 | CGG – 618<br>TGG – 5 | CGG – 607<br>TGG – 3 | CGG – 523<br>TGG – 6 | CGG – 471<br>TGG – 5 | CGG – 424<br>TGG – 3 |
| <u>D796Y</u> | 23948G>T | 23948<br>GAT>TAT | GAT – 16<br>TAT – 70 | GAT – 1155<br>TAT – 0 | GAT – 1409<br>TAT – 5 | GAT – 1313<br>TAT – 10 | GAT – 38<br>TAT – 1257 | GAT – 6<br>TAT – 1438 | GAT – 12<br>TAT – 1223 |
| <u>A1078V</u> | 24795C>T | 24794<br>GCT>GTT | GCT – 219<br>GTT – 3 | GCT – 772<br>GTT - 358 | GCT – 1319<br>GTT – 18 | GCT – 1270<br>GTT – 45 | GCT – 1418<br>GTT – 85 | GCT – 1418<br>GTT – 36 | GCT – 1264<br>GTT – 29 |

\*Only deletions where the adjacent codon was preserved were counted.

**Table S4:** Characteristics of Pfizer BNT162b2 vaccinated participants.

| **Cohort ID** | **Age range (y)** | **Sex** | **Days post-second dose** |
| --- | --- | --- | --- |
| 136023 | 60-69 | M | 11 |
| 136024 | 60-69 | M | 10 |
| 136025 | 50-59 | M | 18 |
| 136026 | 40-49 | F | 9 |
| 136028 | 60-69 | M | 10 |
| 136029 | 20-29 | F | 8 |
| 136071 | 60-69 | M | 128 |
| 136072 | 30-39 | F | 134 |
| 136074 | 20-29 | M | 131 |
| 136075 | 50-59 | M | 152 |
| 136076 | 30-39 | F | 153 |
| 136078 | 30-39 | M | 158 |

## References

1. Khoury DS, Cromer D, Reynaldi A, Schlub TE, Wheatley AK, Juno JA, Subbarao K, Kent SJ, Triccas JA, Davenport MP. Neutralizing antibody levels are highly predictive of immune protection from symptomatic SARS-CoV-2 infection. Nature medicine. 2021:1–7.

2. Earle KA, Ambrosino DM, Fiore-Gartland A, Goldblatt D, Gilbert PB, Siber GR, Dull P, Plotkin SA. Evidence for antibody as a protective correlate for COVID-19 vaccines. Vaccine. 2021;39(32):4423–8.

3. Madhi SA, Baillie V, Cutland CL, Voysey M, Koen AL, Fairlie L, Padayachee SD, Dheda K, Barnabas SL, Bhorat QE, Briner C, Kwatra G, Ahmed K, Aley P, Bhikha S, Bhiman JN, Bhorat AE, du Plessis J, Esmail A, Groenewald M, Horne E, Hwa SH, Jose A, Lambe T, Laubscher M, Malahleha M, Masenya M, Masilela M, McKenzie S, Molapo K, Moultrie A, Oelofse S, Patel F, Pillay S, Rhead S, Rodel H, Rossouw L, Taoushanis C, Tegally H, Thombrayil A, van Eck S, Wibmer CK, Durham NM, Kelly EJ, Villafana TL, Gilbert S, Pollard AJ, de Oliveira T, Moore PL, Sigal A, Izu A, Group N-S, Wits VCG. Efficacy of the ChAdOx1 nCoV-19 Covid-19 Vaccine against the B.1.351 Variant. N Engl J Med. 2021;384(20):1885–98.

4. Garcia-Beltran WF, Lam EC, Denis KS, Nitido AD, Garcia ZH, Hauser BM, Feldman J, Pavlovic MN, Gregory DJ, Poznansky MC. Multiple SARS-CoV-2 variants escape neutralization by vaccine-induced humoral immunity. Cell. 2021;184(9):2372–83. e9.

5. Supasa P, Zhou D, Dejnirattisai W, Liu C, Mentzer AJ, Ginn HM, Zhao Y, Duyvesteyn HME, Nutalai R, Tuekprakhon A, Wang B, Paesen GC, Slon-Campos J, Lopez-Camacho C, Hallis B, Coombes N, Bewley KR, Charlton S, Walter TS, Barnes E, Dunachie SJ, Skelly D, Lumley SF, Baker N, Shaik I, Humphries HE, Godwin K, Gent N, Sienkiewicz A, Dold C, Levin R, Dong T, Pollard AJ, Knight JC, Klenerman P, Crook D, Lambe T, Clutterbuck E, Bibi S, Flaxman A, Bittaye M, Belij-Rammerstorfer S, Gilbert S, Hall DR, Williams MA, Paterson NG, James W, Carroll MW, Fry EE, Mongkolsapaya J, Ren J, Stuart DI, Screaton GR. Reduced neutralization of SARS-CoV-2 B.1.1.7 variant by convalescent and vaccine sera. Cell. 2021;184(8):2201–11 e7.

6. Planas D, Bruel T, Grzelak L, Guivel-Benhassine F, Staropoli I, Porrot F, Planchais C, Buchrieser J, Rajah MM, Bishop E, Albert M, Donati F, Prot M, Behillil S, Enouf V, Maquart M, Smati-Lafarge M, Varon E, Schortgen F, Yahyaoui L, Gonzalez M, De Seze J, Pere H, Veyer D, Seve A, Simon-Loriere E, Fafi-Kremer S, Stefic K, Mouquet H, Hocqueloux L, van der Werf S, Prazuck T, Schwartz O. Sensitivity of infectious SARS-CoV-2 B.1.1.7 and B.1.351 variants to neutralizing antibodies. Nat Med. 2021;27(5):917–24.

7. Wang P, Nair MS, Liu L, Iketani S, Luo Y, Guo Y, Wang M, Yu J, Zhang B, Kwong PD, Graham BS, Mascola JR, Chang JY, Yin MT, Sobieszczyk M, Kyratsous CA, Shapiro L, Sheng Z, Huang Y, Ho DD. Antibody resistance of SARS-CoV-2 variants B.1.351 and B.1.1.7. Nature. 2021;593(7857):130–5.

8. Tegally H, Wilkinson E, Giovanetti M, Iranzadeh A, Fonseca V, Giandhari J, Doolabh D, Pillay S, San EJ, Msomi N, Mlisana K, von Gottberg A, Walaza S, Allam M, Ismail A, Mohale T, Glass AJ, Engelbrecht S, Van Zyl G, Preiser W, Petruccione F, Sigal A, Hardie D, Marais G, Hsiao NY, Korsman S, Davies MA, Tyers L, Mudau I, York D, Maslo C, Goedhals D, Abrahams S, Laguda-Akingba O, Alisoltani-Dehkordi A, Godzik A, Wibmer CK, Sewell BT, Lourenco J, Alcantara LCJ, Kosakovsky Pond SL, Weaver S, Martin D, Lessells RJ, Bhiman JN, Williamson C, de Oliveira T. Detection of a SARS-CoV-2 variant of concern in South Africa. Nature. 2021;592(7854):438–43.

9. Cele S, Gazy I, Jackson L, Hwa SH, Tegally H, Lustig G, Giandhari J, Pillay S, Wilkinson E, Naidoo Y, Karim F, Ganga Y, Khan K, Bernstein M, Balazs AB, Gosnell BI, Hanekom W, Moosa MS, Network for Genomic Surveillance in South A, Team C-K, Lessells RJ, de Oliveira T, Sigal A. Escape of SARS-CoV-2 501Y.V2 from neutralization by convalescent plasma. Nature. 2021;593(7857):142–6.

10. Wibmer CK, Ayres F, Hermanus T, Madzivhandila M, Kgagudi P, Oosthuysen B, Lambson BE, de Oliveira T, Vermeulen M, van der Berg K, Rossouw T, Boswell M, Ueckermann V, Meiring S, von Gottberg A, Cohen C, Morris L, Bhiman JN, Moore PL. SARS-CoV-2 501Y.V2 escapes neutralization by South African COVID-19 donor plasma. Nat Med. 2021;27(4):622–5.

11. Zhou D, Dejnirattisai W, Supasa P, Liu C, Mentzer AJ, Ginn HM, Zhao Y, Duyvesteyn HME, Tuekprakhon A, Nutalai R, Wang B, Paesen GC, Lopez-Camacho C, Slon-Campos J, Hallis B, Coombes N, Bewley K, Charlton S, Walter TS, Skelly D, Lumley SF, Dold C, Levin R, Dong T, Pollard AJ, Knight JC, Crook D, Lambe T, Clutterbuck E, Bibi S, Flaxman A, Bittaye M, Belij-Rammerstorfer S, Gilbert S, James W, Carroll MW, Klenerman P, Barnes E, Dunachie SJ, Fry EE, Mongkolsapaya J, Ren J, Stuart DI, Screaton GR. Evidence of escape of SARS-CoV-2 variant B.1.351 from natural and vaccine-induced sera. Cell. 2021;184(9):2348–61 e6.

12. Wang Z, Schmidt F, Weisblum Y, Muecksch F, Barnes CO, Finkin S, Schaefer-Babajew D, Cipolla M, Gaebler C, Lieberman JA, Oliveira TY, Yang Z, Abernathy ME, Huey-Tubman KE, Hurley A, Turroja M, West KA, Gordon K, Millard KG, Ramos V, Da Silva J, Xu J, Colbert RA, Patel R, Dizon J, Unson-O’Brien C, Shimeliovich I, Gazumyan A, Caskey M, Bjorkman PJ, Casellas R, Hatziioannou T, Bieniasz PD, Nussenzweig MC. mRNA vaccine-elicited antibodies to SARS-CoV-2 and circulating variants. Nature. 2021;592(7855):616–22.

13. Dejnirattisai W, Zhou D, Supasa P, Liu C, Mentzer AJ, Ginn HM, Zhao Y, Duyvesteyn HME, Tuekprakhon A, Nutalai R, Wang B, Lopez-Camacho C, Slon-Campos J, Walter TS, Skelly D, Costa Clemens SA, Naveca FG, Nascimento V, Nascimento F, Fernandes da Costa C, Resende PC, Pauvolid-Correa A, Siqueira MM, Dold C, Levin R, Dong T, Pollard AJ, Knight JC, Crook D, Lambe T, Clutterbuck E, Bibi S, Flaxman A, Bittaye M, Belij-Rammerstorfer S, Gilbert SC, Carroll MW, Klenerman P, Barnes E, Dunachie SJ, Paterson NG, Williams MA, Hall DR, Hulswit RJG, Bowden TA, Fry EE, Mongkolsapaya J, Ren J, Stuart DI, Screaton GR. Antibody evasion by the P.1 strain of SARS-CoV-2. Cell. 2021;184(11):2939–54 e9.

14. Acevedo ML, Alonso-Palomares L, Bustamante A, Gaggero A, Paredes F, Cortés CP, Valiente-Echeverría F, Soto-Rifo R. Infectivity and immune escape of the new SARS-CoV-2 variant of interest Lambda. medRxiv. 2021:2021.06.28.21259673.

15. Uriu K, Kimura I, Shirakawa K, Takaori-Kondo A, Nakada TA, Kaneda A, Nakagawa S, Sato K, Genotype to Phenotype Japan C. Neutralization of the SARS-CoV-2 Mu Variant by Convalescent and Vaccine Serum. N Engl J Med. 2021.

16. Mlcochova P, Kemp S, Dhar MS, Papa G, Meng B, Ferreira I, Datir R, Collier DA, Albecka A, Singh S, Pandey R, Brown J, Zhou J, Goonawardane N, Mishra S, Whittaker C, Mellan T, Marwal R, Datta M, Sengupta S, Ponnusamy K, Radhakrishnan VS, Abdullahi A, Charles O, Chattopadhyay P, Devi P, Caputo D, Peacock T, Wattal DC, Goel N, Satwik A, Vaishya R, Agarwal M, Indian S-C-GC, Genotype to Phenotype Japan C, Collaboration C-NBC-, Mavousian A, Lee JH, Bassi J, Silacci-Fegni C, Saliba C, Pinto D, Irie T, Yoshida I, Hamilton WL, Sato K, Bhatt S, Flaxman S, James LC, Corti D, Piccoli L, Barclay WS, Rakshit P, Agrawal A, Gupta RK. SARS-CoV-2 B.1.617.2 Delta variant replication and immune evasion. Nature. 2021.

17. Liu C, Ginn HM, Dejnirattisai W, Supasa P, Wang B, Tuekprakhon A, Nutalai R, Zhou D, Mentzer AJ, Zhao Y, Duyvesteyn HME, Lopez-Camacho C, Slon-Campos J, Walter TS, Skelly D, Johnson SA, Ritter TG, Mason C, Costa Clemens SA, Gomes Naveca F, Nascimento V, Nascimento F, Fernandes da Costa C, Resende PC, Pauvolid-Correa A, Siqueira MM, Dold C, Temperton N, Dong T, Pollard AJ, Knight JC, Crook D, Lambe T, Clutterbuck E, Bibi S, Flaxman A, Bittaye M, Belij-Rammerstorfer S, Gilbert SC, Malik T, Carroll MW, Klenerman P, Barnes E, Dunachie SJ, Baillie V, Serafin N, Ditse Z, Da Silva K, Paterson NG, Williams MA, Hall DR, Madhi S, Nunes MC, Goulder P, Fry EE, Mongkolsapaya J, Ren J, Stuart DI, Screaton GR. Reduced neutralization of SARS-CoV-2 B.1.617 by vaccine and convalescent serum. Cell. 2021;184(16):4220–36 e13.

18. Barnes CO, Jette CA, Abernathy ME, Dam KA, Esswein SR, Gristick HB, Malyutin AG, Sharaf NG, Huey-Tubman KE, Lee YE, Robbiani DF, Nussenzweig MC, West AP, Jr., Bjorkman PJ. SARS-CoV-2 neutralizing antibody structures inform therapeutic strategies. Nature. 2020;588(7839):682–7.

19. Greaney AJ, Loes AN, Crawford KH, Starr TN, Malone KD, Chu HY, Bloom JD. Comprehensive mapping of mutations in the SARS-CoV-2 receptor-binding domain that affect recognition by polyclonal human plasma antibodies. Cell host & microbe. 2021;29(3):463–76. e6.

20. Greaney AJ, Starr TN, Barnes CO, Weisblum Y, Schmidt F, Caskey M, Gaebler C, Cho A, Agudelo M, Finkin S, Wang Z, Poston D, Muecksch F, Hatziioannou T, Bieniasz PD, Robbiani DF, Nussenzweig MC, Bjorkman PJ, Bloom JD. Mapping mutations to the SARS-CoV-2 RBD that escape binding by different classes of antibodies. Nature Communications. 2021;12(1):4196.

21. McCallum M, De Marco A, Lempp FA, Tortorici MA, Pinto D, Walls AC, Beltramello M, Chen A, Liu Z, Zatta F, Zepeda S, di Iulio J, Bowen JE, Montiel-Ruiz M, Zhou J, Rosen LE, Bianchi S, Guarino B, Fregni CS, Abdelnabi R, Foo SC, Rothlauf PW, Bloyet LM, Benigni F, Cameroni E, Neyts J, Riva A, Snell G, Telenti A, Whelan SPJ, Virgin HW, Corti D, Pizzuto MS, Veesler D. N-terminal domain antigenic mapping reveals a site of vulnerability for SARS-CoV-2. Cell. 2021;184(9):2332–47 e16.

22. Cerutti G, Guo Y, Zhou T, Gorman J, Lee M, Rapp M, Reddem ER, Yu J, Bahna F, Bimela J. Potent SARS-CoV-2 neutralizing antibodies directed against spike N-terminal domain target a single supersite. Cell Host & Microbe. 2021;29(5):819–33. e7.

23. Suryadevara N, Shrihari S, Gilchuk P, VanBlargan LA, Binshtein E, Zost SJ, Nargi RS, Sutton RE, Winkler ES, Chen EC. Neutralizing and protective human monoclonal antibodies recognizing the N-terminal domain of the SARS-CoV-2 spike protein. Cell. 2021;184(9):2316–31. e15.

24. Chi X, Yan R, Zhang J, Zhang G, Zhang Y, Hao M, Zhang Z, Fan P, Dong Y, Yang Y. A neutralizing human antibody binds to the N-terminal domain of the Spike protein of SARS-CoV-2. Science. 2020;369(6504):650–5.

25. Karim F, Gazy I, Cele S, Zungu Y, Krause R, Bernstein M, Khan K, Ganga Y, Rodel HE, Mthabela N, Mazibuko M, Muema D, Ramjit D, Ndung’u T, Hanekom W, Gosnell B, Team C-K, Lessells RJ, Wong EB, de Oliveira T, Moosa Y, Lustig G, Leslie A, Kloverpris H, Sigal A. HIV status alters disease severity and immune cell responses in beta variant SARS-CoV-2 infection wave. Elife. 2021;10.

26. Avelino-Silva VI, Miyaji KT, Mathias A, Costa DA, de Carvalho Dias JZ, Lima SB, Simoes M, Freire MS, Caiaffa-Filho HH, Hong MA. CD4/CD8 ratio predicts yellow fever vaccine-induced antibody titers in virologically suppressed HIV-infected patients. JAIDS Journal of Acquired Immune Deficiency Syndromes. 2016;71(2):189–95.

27. Ho DD, Neumann AU, Perelson AS, Chen W, Leonard JM, Markowitz M. Rapid turnover of plasma virions and CD4 lymphocytes in HIV-1 infection. Nature. 1995;373(6510):123–6.

28. Murphy EL, Collier AC, Kalish LA, Assmann SF, Para MF, Flanigan TP, Kumar PN, Mintz L, Wallach FR, Nemo GJ. Highly active antiretroviral therapy decreases mortality and morbidity in patients with advanced HIV disease. Annals of internal medicine. 2001;135(1):17–26.

29. Hoffman SA, Costales C, Sahoo MK, Palanisamy S, Yamamoto F, Huang C, Verghese M, Solis DC, Sibai M, Subramanian A. SARS-CoV-2 Neutralization Resistance Mutations in Patient with HIV/AIDS, California, USA. Emerging Infectious Diseases. 2021;27(10).

30. Karim F, Moosa MY, Gosnell B, Sandile C, Giandhari J, Pillay S, Tegally H, Wilkinson E, San EJ, Msomi N. Persistent SARS-CoV-2 infection and intra-host evolution in association with advanced HIV infection. medRxiv. 2021.

31. Chen L, Zody MC, Di Germanio C, Martinelli R, Mediavilla JR, Cunningham MH, Composto K, Chow KF, Kordalewska M, Corvelo A, Oschwald DM, Fennessey S, Zetkulic M, Dar S, Kramer Y, Mathema B, Germer S, Stone M, Simmons G, Busch MP, Maniatis T, Perlin DS, Kreiswirth BN. Emergence of Multiple SARS-CoV-2 Antibody Escape Variants in an Immunocompromised Host Undergoing Convalescent Plasma Treatment. mSphere. 2021;6(4):e0048021.

32. Choi B, Choudhary MC, Regan J, Sparks JA, Padera RF, Qiu X, Solomon IH, Kuo HH, Boucau J, Bowman K, Adhikari UD, Winkler ML, Mueller AA, Hsu TY, Desjardins M, Baden LR, Chan BT, Walker BD, Lichterfeld M, Brigl M, Kwon DS, Kanjilal S, Richardson ET, Jonsson AH, Alter G, Barczak AK, Hanage WP, Yu XG, Gaiha GD, Seaman MS, Cernadas M, Li JZ. Persistence and Evolution of SARS-CoV-2 in an Immunocompromised Host. N Engl J Med. 2020;383(23):2291–3.

33. Clark SA, Clark LE, Pan J, Coscia A, McKay LGA, Shankar S, Johnson RI, Brusic V, Choudhary MC, Regan J, Li JZ, Griffiths A, Abraham J. SARS-CoV-2 evolution in an immunocompromised host reveals shared neutralization escape mechanisms. Cell. 2021;184(10):2605–17 e18.

34. Kemp SA, Collier DA, Datir RP, Ferreira I, Gayed S, Jahun A, Hosmillo M, Rees-Spear C, Mlcochova P, Lumb IU, Roberts DJ, Chandra A, Temperton N, Collaboration C-NBC-, Consortium C-GU, Sharrocks K, Blane E, Modis Y, Leigh KE, Briggs JAG, van Gils MJ, Smith KGC, Bradley JR, Smith C, Doffinger R, Ceron-Gutierrez L, Barcenas-Morales G, Pollock DD, Goldstein RA, Smielewska A, Skittrall JP, Gouliouris T, Goodfellow IG, Gkrania-Klotsas E, Illingworth CJR, McCoy LE, Gupta RK. SARS-CoV-2 evolution during treatment of chronic infection. Nature. 2021;592(7853):277–82.

35. Weigang S, Fuchs J, Zimmer G, Schnepf D, Kern L, Beer J, Luxenburger H, Ankerhold J, Falcone V, Kemming J, Hofmann M, Thimme R, Neumann-Haefelin C, Ulferts S, Grosse R, Hornuss D, Tanriver Y, Rieg S, Wagner D, Huzly D, Schwemmle M, Panning M, Kochs G. Within-host evolution of SARS-CoV-2 in an immunosuppressed COVID-19 patient as a source of immune escape variants. Nat Commun. 2021;12(1):6405.

36. Johnson BA, Xie X, Bailey AL, Kalveram B, Lokugamage KG, Muruato A, Zou J, Zhang X, Juelich T, Smith JK, Zhang L, Bopp N, Schindewolf C, Vu M, Vanderheiden A, Winkler ES, Swetnam D, Plante JA, Aguilar P, Plante KS, Popov V, Lee B, Weaver SC, Suthar MS, Routh AL, Ren P, Ku Z, An Z, Debbink K, Diamond MS, Shi P-Y, Freiberg AN, Menachery VD. Loss of furin cleavage site attenuates SARS-CoV-2 pathogenesis. Nature. 2021;591(7849):293–9.

37. Greaney AJ, Starr TN, Eguia RT, Loes AN, Khan K, Karim F, Cele S, Bowen JE, Logue JK, Corti D, Veesler D, Chu HY, Sigal A, Bloom JD. A SARS-CoV-2 variant elicits an antibody response with a shifted immunodominance hierarchy. bioRxiv. 2021:2021.10.12.464114.

38. On YB, Noor E, Gottlieb N, Sigal A, Milo R. The importance of time post-vaccination in determining the decrease in vaccine efficacy against SARS-CoV-2 variants of concern. medRxiv. 2021:2021.06.06.21258429.

39. Smith DJ, Lapedes AS, De Jong JC, Bestebroer TM, Rimmelzwaan GF, Osterhaus AD, Fouchier RA. Mapping the antigenic and genetic evolution of influenza virus. science. 2004;305(5682):371–6.

40. Korber B, Muldoon M, Theiler J, Gao F, Gupta R, Lapedes A, Hahn B, Wolinsky S, Bhattacharya T. Timing the ancestor of the HIV-1 pandemic strains. science. 2000;288(5472):1789–96.

41. Minor PD, Ferguson M, Evans DM, Almond JW, Icenogle JP. Antigenic structure of polioviruses of serotypes 1, 2 and 3. Journal of General Virology. 1986;67(7):1283–91.

42. Guzman MG, Alvarez M, Rodriguez-Roche R, Bernardo L, Montes T, Vazquez S, Morier L, Alvarez A, Gould EA, Kourí G. Neutralizing antibodies after infection with dengue 1 virus. Emerging infectious diseases. 2007;13(2):282.

43. Meintjes G, Stek C, Blumenthal L, Thienemann F, Schutz C, Buyze J, Ravinetto R, van Loen H, Nair A, Jackson A. Prednisone for the prevention of paradoxical tuberculosis-associated IRIS. New England Journal of Medicine. 2018;379(20):1915–25.

44. Khan K, Lustig G, Bernstein M, Archary D, Cele S, Karim F, Smith M, Ganga Y, Jule Z, Reedoy K, Miya Y, Mthabela N, Team C-K, Lessells R, de Oliveira T, Gosnell BI, Karim SA, Garrett N, Hanekom W, Bekker LG, Gray G, Blackburn JM, Moosa M-YS, Sigal A. Immunogenicity of SARS-CoV-2 infection and Ad26.CoV2.S vaccination in people living with HIV. medRxiv. 2021:2021.10.08.21264519.

45. Sykes W, Mhlanga L, Swanevelder R, Glatt TN, Grebe E, Coleman C, Pieterson N, Cable R, Welte A, van den Berg K. Prevalence of anti-SARS-CoV-2 antibodies among blood donors in Northern Cape, KwaZulu-Natal, Eastern Cape, and Free State provinces of South Africa in January 2021. Research Square. 2021.

